# Mathematically modeling worried-well behavior during infectious disease outbreaks

**DOI:** 10.1101/2025.02.06.25321784

**Authors:** Bismark Singh, Dmitry Gromov

## Abstract

Mitigation and control of novel infectious disease outbreaks requires a strategic response from public health officials. Such a response requires curtailment of both the underlying pathogen as well as public concern. There is an extensive amount of literature on the former, however empirical models catered to the latter are lacking. We seek to fill this gap by presenting a mathematical model that governs spread of “worried-well” behavior within an infectious disease outbreak. Such behavioral concerns, that are marked by an absence of the underlying pathogen infection, manifest themselves in varied degrees ranging from protesting behavior against public health measures to overly isolating behavior leading to mental anxiety. Our work shows different strategies are required for different degrees of worried-well behavior. We provide a framework to mathematically formulate such behavior and provide guidance on controlling surges of such population groups under varied proportions. Since compliance to public health measures is strongly influenced by effective public health messaging, estimates of such population groups are imperative to public health officials.

## Introduction

Novel infectious disease outbreaks, that arise from new strains that are previously unidentified in humans, are characterized by uncertainty. Such uncertainty manifests itself in terms of the spread of the inducing pathogen as well as the spread of associated public opinion and perceptions of the outbreak [1]. There is a significant body of epidemiological literature devoted to the former that estimates trajectories of both symptomatic and asymptomatic pathogen-infected individuals, see, e.g., [2]. However, there is little quantitative research on how behavioral changes (e.g., worry or concern) spread during a pathogen-induced pandemic. An adequate understanding of the dynamics of such behavioral changes is imperative for pandemic preparedness and control for two key reasons.

First, public perception of the pandemic drives compliance towards or away from disease control policies. The resulting behavioral changes — even in uninfected individuals — directly lead to an increased or decreased overall spread of the underlying pathogen; see, e.g., [3, 4]. An example of such a behavioral change is social distancing. Significant amount of evidence from the COVID-19 pandemic showed that following mandates for social distancing reduces the spread of the pathogen [5]. However, compliance to social distancing mandates was determined by an individual’s perception of the risk of the outbreak [6]. Some individuals became fearful and self-protective anxiously avoiding other individuals [7]. Other individuals lost their trust in governmental public health services and actively protesting against the COVID-19 mitigation strategies [8]. Both these extreme behavioral policies require a different pandemic response strategy from a policymaker for mitigation of the disease spread.

Second, asymptomatic testing is often unavailable or unreliable at the start of the pandemic [9]. During this period, public healthcare policy of allocating scarce resources is driven by professional judgment [10, 11] which is not always able to correctly identify a genuinely sick individual from one who is worried or concerned about the new outbreak [1]. Thus, scarce therapeutic resources risk being consumed by uninfected populations leading to shortages of resources for the pathogen-infected populations. Ultimately, this could lead to an increased spread of the pathogen if a suitable vaccine is not identified within a reasonable amount of time. Thus, good estimates of counts of concerned individuals are required to correctly identify demand for resources which ultimately help determining fair and effective resource allocation policies.

Beginning from a slight trace, such behavioral concerns can quickly overwhelm a healthcare system by frequent medical visits for diagnosis and assurance. A poll conducted in February 2020 showed that, although only four Canadians were infected by COVID-19, 2.6 million people were very concerned [1]. Such behavior is typically without malicious intent; however, it could increase the spread of the disease by the concerned individual’s frequent hospital visits thereby compounding the challenge for public health officials. Indeed, counts of such individuals could be twenty times the genuinely infected population making them among the hardest aspects of public-health handling during outbreaks [12]. This challenge is particularly difficult to address during the early days of a novel outbreak when there is a larger degree of uncertainty about both the *perceived* and *actual* risk of the disease. Perceived risk of the outbreak directly influences preventative behavior [13]. Thus, accurate estimation of the perceived risk is critical in determining the counts of such populations. As we mention above, in the absence of such data-driven guidance, policymakers who are more prone to taking risk (“risk-taking”) could erroneously assume low counts of such populations resulting in an overwhelmed healthcare system, while policymakers who are less prone to taking risk (“risk-averse”) might budget too many resources for an outbreak of a low-scale leading to economic losses. Former US president Franklin Roosevelt’s decision to substantially increase healthcare funding in response to the polio pandemic and Chinese president Xi Jinping’s decision to enforce strict lockdowns in response to the COVID-19 pandemic are two examples of risk-taking behavior by politicians. Former German chancellor Angela Merkel’s decision to not significantly alter German healthcare policy during the Ebola outbreak and former Indian prime minister Manmohan Singh’s decision to not aggressively contain the 2009 H1N1 flu are examples of risk-averse behavior by politicians.

In this work, we present a mathematical model that estimates counts of population groups who are uninfected by the pathogen but are still concerned about the disease. The medical literature denotes such populations by the term “worried-well”. They trace their origin to work in the 1970s where patients entering a health maintenance organization (HMO) were divided into three categories: well, asymptomatically sick, and truly sick [14]. Literature on sexually transmitted diseases recognizes such groups prior to this classification; see, e.g., a study of the psychiatric phenomenon “syphilophobia” (derived from syphilis) where patients appear fearful of medical disease yet present no physical symptoms of the disease [15], or a study of populations seeking reassurance in the absence of infection during the Acquired Immunodeficiency Syndrome (AIDS) epidemic in the late 1980s [16]. At an individual level, such concerns arise due to an emotional response; e.g., anxiety, fear, or distress. At a social level, such concern spreads through so-called “mass psychogenic illnesses”; e.g., from a physical trigger such as smell [17, 18], administration of a vaccine [19], or public concern following catastrophic events [20]. Past work classifies such concerned populations into three subgroups based on the nature of their emotional disturbance: (a) individuals experiencing disease symptoms due to anxiety but without pathogen exposure, (b) individuals working in high-risk activities seeking reassurance of their concerns but without any symptoms of the pathogen, and (c) those with anxiety following a traumatic event [21, 22]. Within this work, we categorize individuals as worried-well if they alter their behavior within a pathogen-induced disease outbreak while being uninfected by the underlying pathogen. We consider different degrees of behavioral change, motivated by an individual’s perceived risk, that lead to an increased or decreased level of contacts. With this background, the aim of our work is to develop guidance for healthcare policymakers on how to respond and how to control surges in the worried-well populations during the early days of a pathogen-induced infectious disease outbreak.

The following are the two key contributions of this article.

a. We present a novel mathematical framework to estimate worried-well populations within a pathogen-induced disease outbreak. Further, our framework allows an inclusion of both individuals who protest against the governmental response as well as those who over-enthusiastically embrace it.
b. We provide empirical guidance to a policymaker on how to respond at the start of the pandemic when there is limited information on the spread of both worry and the disease. Specifically, we analyze two policymaking responses of holding onto scarce medical resources or immediately releasing them.s

The rest of this article is organized as follows. In Section, we describe our compartment-based epidemiological model that governs the spread of the disease and estimates counts of the worried-well population. Section presents the main results of this work; here, we analyze our mathematical model and provide guidance for a policymaker to ensure preparedness for different levels of behavioral compliance. We conclude with a summary and limitations of our work in Section.

## Materials and methods

We consider a pathogen-induced infectious disease outbreak, where both worried-well and pathogen-infected populations evolve over time. The spread of worried-well behavior is similar to that of an infection, and both processes propagate independently via contacts with the correspondingly infected individuals. However, unlike a pathogen that transmits by physical proximity and/or contact, worry additionally propagates via the spread of information; e.g., public health messaging [23], governmental campaigns [24], propaganda [25], and online social networks [26]. In this sense, we formulate a model similar to the basic two-disease model of Blyuss and Kyrychko [27]. Table 1 summarizes the notation we employ.

**Table 1.** Notation for compartmental model (1).

| Parameter | Description | Units |
| --- | --- | --- |
| Compartments |  |  |
| $S$ | fraction of individuals susceptible to both pathogen and worry | |
| $I_P$ | fraction of individuals infected by a pathogen who may or may not be worried (i.e., genuinely sick) | |
| $I_W$ | fraction of individuals worried but not infected by a pathogen (i.e., worried-well) | |
| $R_P$ | fraction of individuals recovered from the pathogen infection | |
| Rates |  |  |
| $\alpha$ | behavioral modifier for the worried-well | $\alpha \in \mathbb{R}^+$ |
| $\beta_P$ | transmission coefficient of pathogen | $[\text{time}^{-1}]$ |
| $\beta_W$ | transmission coefficient of worried-well | $[\text{time}^{-1}]$ |
| $\beta_{WP}$ | transmission coefficient of worry from contact with a sick individual | $[\text{time}^{-1}]$ |
| $\gamma_P$ | transition rate of recovery from pathogen | $[\text{time}^{-1}]$ |
| $\gamma_W$ | transition rate of recovery from worry | $[\text{time}^{-1}]$ |
| $\delta_P$ | transition rate at which individuals recovered from pathogen become susceptible again | $[\text{time}^{-1}]$ |

We divide the host population, *N*, into three groups of compartments: susceptible (*S*), infected (*I*), and recovered (*R*). We assume a negligible mortality of the underlying pathogen-induced disease and a short time interval which allows us to ignore changes to the demographics; thus, the total population *N* remains constant. Individuals in compartment *S* are susceptible to both worry and the pathogen. We consider two compartments for the infected individuals: the genuinely sick population that is infected by the pathogen (*I*_*P*_), and the worried-well population (*I*_*W*_). Individuals in the compartment *I*_*W*_ are not infected by the pathogen but are still worried, and it is an estimate of these individuals that is central to our work. We include individuals who are infected by the pathogen and are also worried about their condition within the compartment *I*_*P*_; i.e., this compartment includes genuinely sick populations irrespective of whether they are worried or not. Thus, we consider these concerns are genuinely distinguished from the concerns of the worried-well individuals. We do so since infected individuals change their behavior and lifestyles during a disease outbreak; see, e.g., [28, 29]. Pathogen-infected individuals in compartment *I*_*P*_ recover following either a pharmaceutical intervention (e.g., antivirals or antibiotics), a period of social-distancing, or rest. With their recovery from the pathogen, their immediate worry of re-infection also temporarily ceases. Hence, we assume that recovery from the pathogen confers a temporary immunity both to the pathogen and to worry. The recovered individuals enter the compartment *R*_*P*_. On the other hand, worry, when taken alone, does not develop an immunity. Thus, individuals, who recover from worry may get worried again. Here, we are motivated by previous studies that demonstrate recurrence of anxiety disorders [30]

The spread of pathogen is governed by contacts with individuals in the *I*_*P*_ compartment alone. However, worried-well behavior spreads via contacts of two types: those with individuals in the compartment *I*_*P*_, and those with individuals in the compartment *I*_*W*_. Thus, as we present in Fig. 1, individuals enter the *I*_*P*_ compartment from both the *S* and *I*_*W*_ compartments, while they enter the *I*_*W*_ compartment from *S* alone. We denote the transmission coefficients for these three types of contacts by *β*_*P*_, *β*_*W*_, and *β*_*W P*_, respectively. We then have the two forces of infection (see, e.g., [31]) defined as follows: *λ*_*P*_ = *β*_*P*_ *I*_*P*_ and *λ*_*W*_ = *β*_*W P*_ *I*_*P*_ + *β*_*W*_ *I*_*W*_. Analogously, we let *γ*_*P*_ and *γ*_*W*_ denote the transition rates corresponding to the removal of the pathogen and worry, respectively. These transition rates equal the inverse average duration of the respective conditions, while *δ*_*P*_ is the inverse average duration of the immunity to the pathogen.

**Fig 1.**
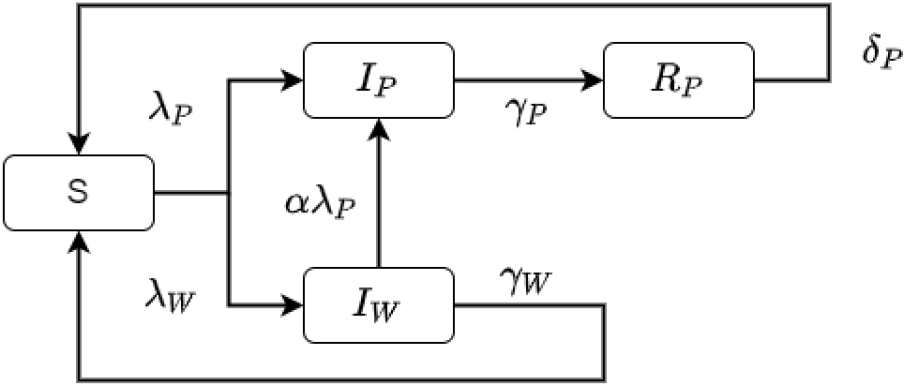
Block diagram for compartmental model (1).

As we mention in Section, worried-well behavior results in an altered lifestyle for several worried-well individuals. We model such behavioral changes via a behavioral factor, *α >* 0: (a) *cautious* individuals who excessively decrease their contacts have *α <* 1, while (ii) *protesting* individuals who excessively increase their contacts have *α >* 1. The default state where an individual is simply worried but does not alter his/her behavior is modeled via *α* = 1. This behavioral factor changes the force of infection, *λ*_*P*_, from the worried-well to the pathogen-infected compartments.

Since the total population is constant, the model equations are formulated in terms of the fractions of the total population. This implies that all system states belong to the interval [0, 1] and the sum of the variables equals 1, i.e., *S* + *I*_*P*_ + *I*_*W*_ + *R*_*P*_ = 1. With this background, we have the following epidemiological compartmental model.

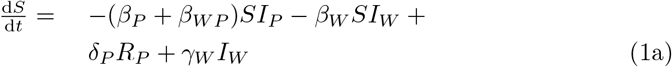

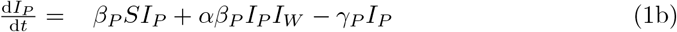

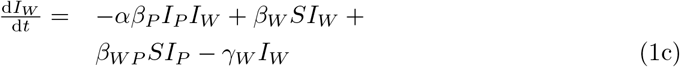

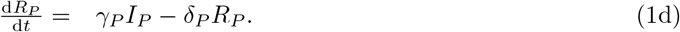

Equations (1a)-(1d) denote the four transitions from the four compartments presented in Fig. 1. Then, the basic reproduction number for model (1) computed using the next-generation matrix method (see, e.g., [32]) equals the maximum of the two respective reproduction numbers (see, e.g., [27]):

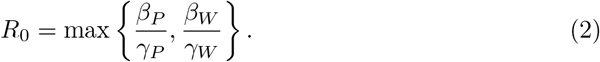

If *R*_0_ *>* 1, disease, worry, or both become endemic in the population. Although the reproduction number is important in determining the expected number of infections, our work is focused on the early-stage transient dynamics of the disease spread that are marked with uncertainty. However, interestingly, the reproduction number is independent of *β*_*W P*_. Intuitively this is so because individuals infected with the pathogen also transmit worry. To further see this, consider the disease-free equilibrium 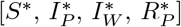 with the corresponding disease free steady state [1, 0, 0, 0]. Then, as described in [27], linearizing the system of equations in model (1) for the three independent variables [*I*_*p*_, *I*_*W*_, *R*_*P*_] near the disease free steady state provides the following matrix (note that *S* is obtained from *S* + *I*_*P*_ + *I*_*W*_ + *R*_*P*_ = 1).

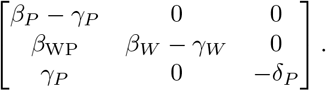

All eigenvalues of this matrix must be negative to ensure stability of the disease-free equilibrium state. The three eigenvalues are *β*_*P*_ *− γ*_*P*_, *β*_*W*_ *− γ*_*W*_ and *−δ*_*P*_. The third eigenvalue is trivially negative, while the requirement of negativity of the first two provides the condition *R*_0_ *<* 1 for the basic reproduction number given by equation (2).

The parameters we use in our numerical experiments are informed by early estimates during the COVID-19 pandemic. We use 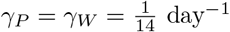 following [33] where the estimated mean time from the onset of symptoms to two negative RT-PCR tests is taken as two weeks. We consider worry to persist equally long, on average, as the pathogen. We consider the basic reproduction number of the pathogen-induced disease,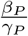, as 10.3 following [34]; then, *β*_*P*_ = 0.74. We consider a conservative estimate of the average duration of immunity to the pathogen as 8 months following [35]; thus,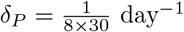. We are unaware of any studies providing estimates of the transmission rate or the basic reproduction number for worry. We consider *β*_*W*_ = *β*_*W P*_ = 0.7; i.e., the reproduction number of the worry-induced disease is 9.8 which is slightly less than the reproduction number of the pathogen-induced disease.

## Results and Analysis

In the absence of asymptomatic testing, the quantity *I*_*P*_ + *I*_*W*_ denotes the total population burden imposed on a healthcare system. This quantity is not necessarily the total individuals seeking scarce therapeutic resources as some groups of the worried-well population, *I*_*W*_, might intentionally not seek resources as we explain below. Individuals without symptoms formed the non-priority group for testing as of March 22, 2020 by the US Centers for Disease Control and Prevention (CDC) [36]. Thus, in the early days of a novel disease outbreak, scarce resources consumed by individuals in the *I*_*W*_ compartment deprive those in the *I*_*P*_ compartment. The fraction 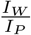 presents one measure of the public pressure presented by the worried-well population on healthcare policymakers.

Resource allocation policies implemented during the phase of the pandemic when this ratio is greater than one (and, asymptomatic testing is unavailable) should respond to reducing the dominating number of worried-well populations. It is in this phase that a strategic policy of holding on to scarce resources, in anticipation of greater demand at a later stage by the genuinely infected population, is advisable since it prevents consumption by the undetectable worried-well populations. On the other hand, resource allocation policies when this ratio is less than one should respond by swiftly combating the spread of the underlying pathogen. In this situation a policymaker can essentially treat the small fractions of worried-well populations similar to the susceptible individuals. Then, a more relaxed policy of immediately allocating all resources (even when resources are scarce and asymptomatic testing is unavailable) is advisable. We analyze these situations below.

We first consider the default case of *α* = 1; see, Fig. 2b. We observe that the peak of the infected population, occurring just before two weeks into the outbreak, is larger than the worried-well population; however, worry dominates the pathogen at the very start of the outbreak. This situation was indeed observed in the early days of the COVID-19 outbreak (in early February 2020); e.g., the large magnitude of worried-well individuals in countries where there were practically no infected populations at that time [1].

**Fig 2.**
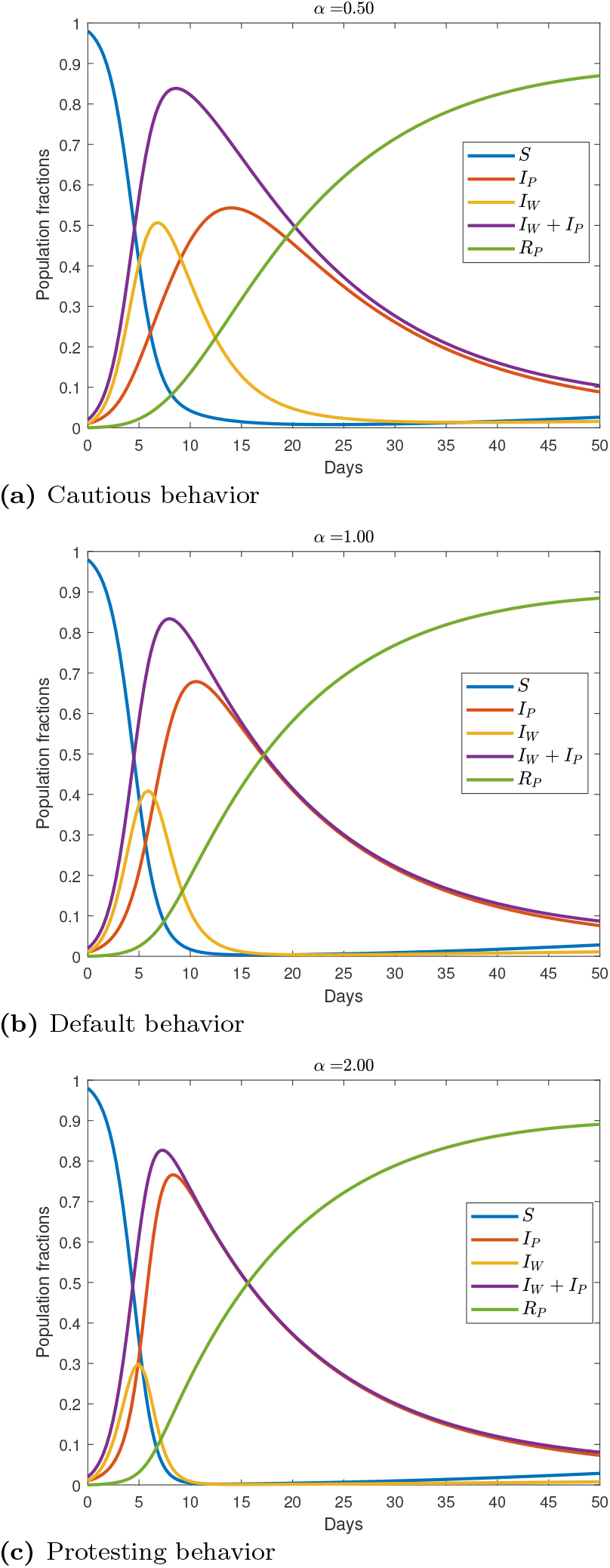
Propagation of pathogen-induced disease and worried-well behavior in three behavioral regimes.

Following this peak, worry starts declining, however the pathogen-infected population continues to rise. This is because both the worried-well and the susceptible populations are getting genuinely infected in addition to waning of the worry; see, model (1). This waning phase also reflects a period of so-called pandemic fatigue where individuals stop worrying about the disease, see, e.g., [37]. We capture this via the parameter *γ*_*W*_. However, mitigating disease spread in both the infected populations requires different intervention strategies. Controlling the spread of a pathogen involves the traditionally well-studied pandemic control measures, such as enforcing face-masks (if available) and social distancing for the general population. Controlling the inflow of worried-well populations into the pool of the infected requires a reduction in the transmission rate of this group alone. We model this latter situation with a cautious and protesting behavior governed by regimes of *α <* 1 and *α >* 1, respectively, that we consider next.

Effective public health messaging and informational campaigns can assist in social compliance by increasing cautious behavior (modeled via *α <* 1) in the uninfected population [3]. For example, past studies show individuals who received news from websites engage in higher levels of social distancing than those receiving news from social media [38]. Fig. 2a presents our results for this situation. The peak of the infected population, *I*_*P*_, in Fig. 2a is lower than that in Fig. 2b even though the overall population seeking resources, *I*_*P*_ + *I*_*W*_, is almost the same in the two situations. Cautious behavior results in significantly larger amount of worried-well populations and lower amount of genuinely infected populations than the default behavior. Here, the worried-well population risks imposing a large amount of public pressure on the policymaker (see, the ratio 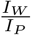 in Fig. 3). Indeed, bipartisan politics influences such pressure; e.g., during the early days of both the Ebola and COVID-19 pandemic, US Republican and Democratic politicians cast blame on each other, respectively, for not doing enough [39]. Further, during the 2009 H1N1 influenza pandemic, consultations by the worried-wells surged in New York City [40].

**Fig 3.**
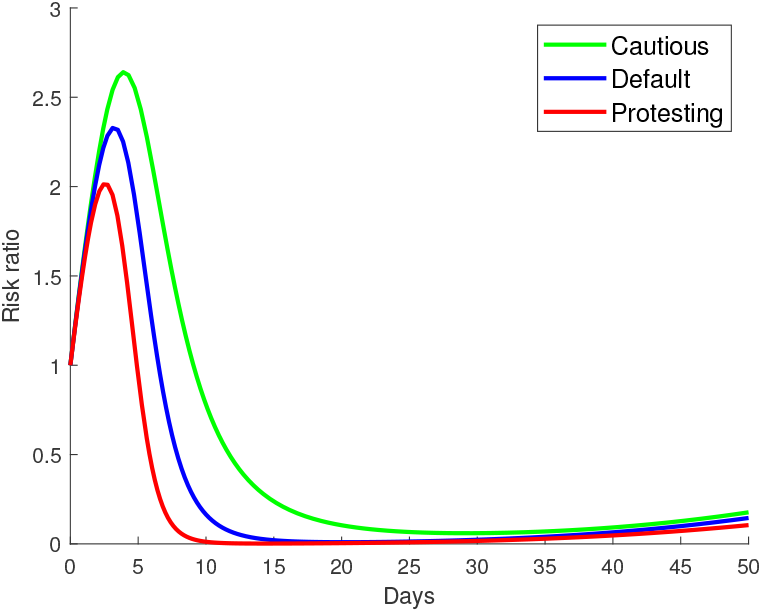
A measure of the policymaker’s pressure during the early days of a disease outbreak given by the ratio of a policymaker’s perceived risk and actual risk (i.e., 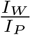), in three behavioral regimes.

Within the cautious regime, the availability of diagnostic asymptomatic testing after the initial phase of high uncertainty and skepticism of the disease spread could ease such public pressure. Then, medical professionals could distinguish between infected and uninfected population groups, thereby realizing that the actual risk (as measured by *I*_*P*_) is lower than the perceived risk (as measured by *I*_*P*_ + *I*_*W*_). At this point, healthcare policies driven by tempering panic and public concerns are advisable (such measures reduce the *I*_*W*_ population by increasing the recovery into the *S* compartment). Examples of such policies include reassuring tweets [41] and holding community rituals [42]. It is important to note that this suggestion does not mean a relaxation of social distancing or adherence measures since the susceptible population is, naturally, vulnerable to get infected.

Now, consider the contrasting regime of *α >* 1 where the worried-well populations display behaviors promoting increased contact-rates. Such behavior manifests itself in the form of increased amount of hospital visits to seek reassurance [22], or protests against lockdown measures [8]. Although both these behaviors are contrasting (the individual seeking reassurance is genuinely concerned, while the individual protesting is belittling the threat), they lead to the same result: an increased contact rate, resulting in an increase in the number of infected individuals. As we show in Fig. 2c, the peak of the infected population, *I*_*P*_, is larger than the other two regimes. However, although the transition of the worried-wells into the infected population is faster in this regime (compared to both the default and cautious behavior), the initial phase is again dominated by worry.

Within this protesting regime, the peak demand for resources (bounded by *I*_*P*_ + *I*_*W*_) arrives sooner than the other two regimes. Such crisis management presents a considerably larger amount of public pressure for the public health official on two grounds. First, because the public health official has had lesser time for preparedness. Second, because the perceived risk is indeed (almost) actual. This is true even in the highly optimistic scenario that asymptomatic testing is available early on since the ratio 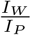 is smaller than the default and cautious regimes; likely, asymptomatic testing will not be available in the initial period due to the disease’s novelty. Within this regime, disease control policies must cater to, both, mitigating the pathogen and addressing the concerns of the worried-wells. An authoritarian policy that enforces a strict lockdown immediately addresses both these challenges together [43]; although, such policies risk, both, larger protests later on [8] as well as feelings of depression and hopelessness in the public [44]. More liberal resource allocation policies that extensively distribute the available scarce resources, without holding them back, are advisable for this regime. This is because the risk of wastage of resources by consumption from the uninfected population is low. Such an allocation policy is also appeasing to worried-well individuals seeking reassurance who might feel their concerns are not simply being dismissed [45]. It is unclear how such a policy would be interpreted by worried-well individuals who protest against governmental policies.

The analysis so far assumes the same amount of worried and genuinely infected individuals (both set to 1% of the total population) to initialize the disease progression. To determine whether our results are biased due to this choice of the initial proportions, we conducted a similar numerical experiment for different initial proportions of the worried and genuinely infected individuals (see, Appendix). We observe that the overall trend remains largely unaffected by this choice; naturally, the severity of the disease (i.e., the number of pathogen-induced or worried-well populations) is affected. For example, even when the initially worried population is ten times less than the initially infected population, the profiles of genuinely infected and worried remain the same (see Fig. 4). Moreover, the peak value of the fraction 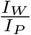 exceeds two (see, Fig. 5 in Appendix). In contrast, this peak value surpasses 2.5 for equal initial proportions of the two groups. Such patterns are explained by the fact that at the onset of the outbreak the basic reproduction numbers of both the infection and worry are relative high (almost 10), which overpower the effects of the initial proportions. On the other hand, we observe that a large proportion of initially worried-well populations increases the progression of worry, even though the overall pattern remains the same (cf. Fig. 4). Specifically, the ratio 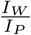 is about 10 for the case when the initial worried population is ten times larger than the population of infected (see Fig. 5). In this sense, we can say that worry has a self-enforcing effect.

**Fig 4.**
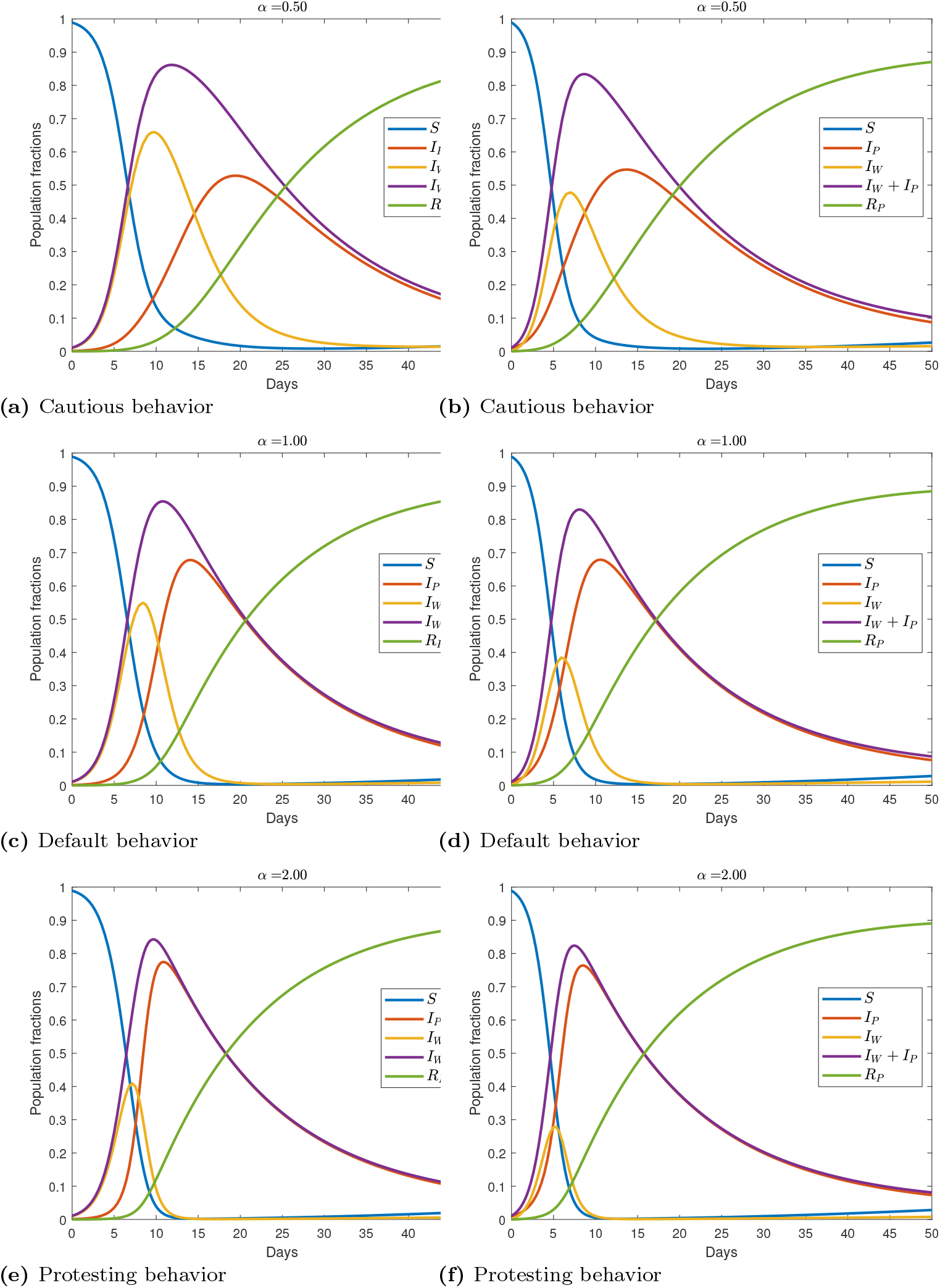
Analogous graphs to Fig. 2 of the main text for the propagation of pathogen-induced disease and worried-well behavior in three behavioral regimes. The left panel denotes the case when the initially infected population is ten times less than the initially worried population. The right panel denotes the case when the initially worried population is ten times less than the initially infected population.

**Fig 5.**
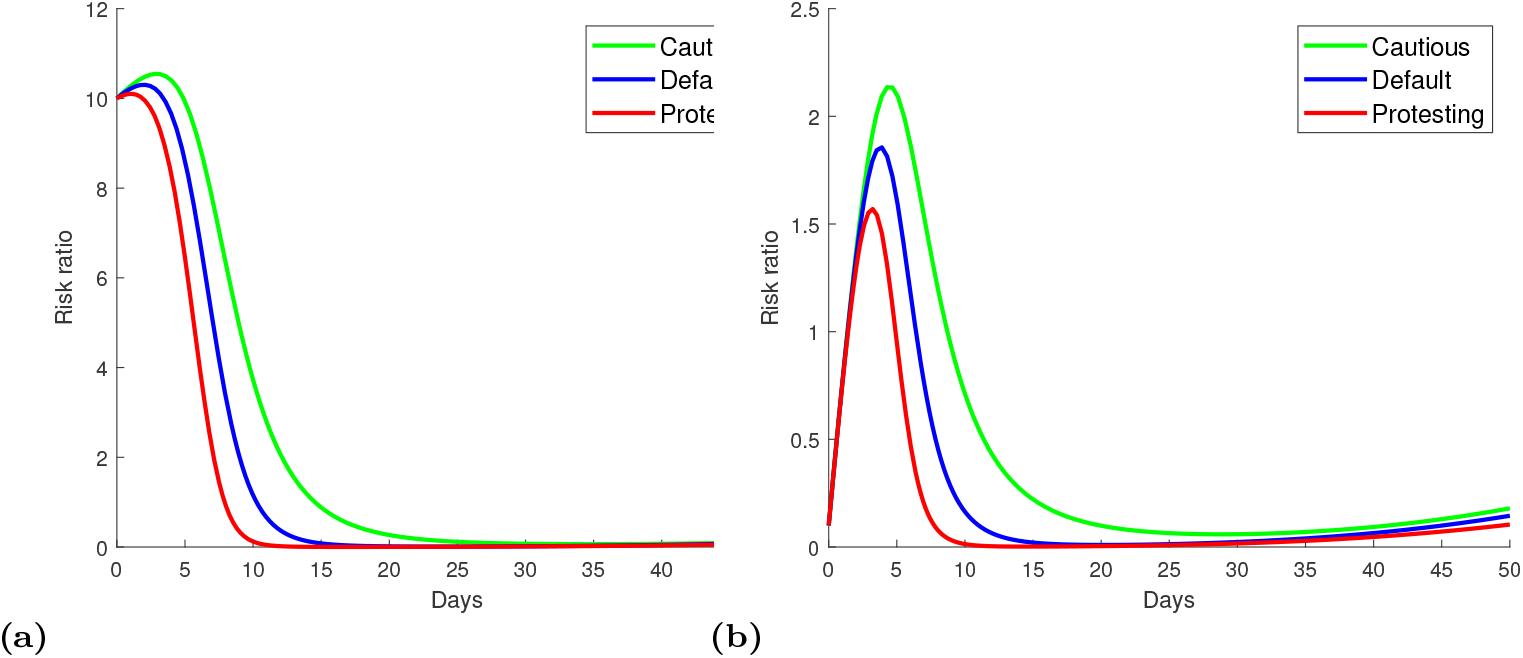
Analogous graphs to Fig. 3 of the main text determining a measure of the policymaker’s pressure during the early days of a disease outbreak given by the ratio of a policymaker’s perceived risk and actual risk (i.e., 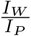), in three behavioral regimes. Fig. 5a and Fig. 5b consider the initially infected population to be ten times less and more than the initially worried population, respectively.

## Conclusions

The World Health Organization (WHO)’s Director–General remarked on February 15, 2020: “We’re not just fighting an epidemic; we’re fighting an infodemic. Fake news spreads faster and more easily than this virus, and is just as dangerous.” [46]. Our analysis in Section provides support of this statement where even small traces of unfounded concerns quickly escalate rising numbers of infected populations. Past research lacks a way to model and empirically estimate counts of such populations, and our research seeks to fill this gap. Knowledge of such estimates allows policymakers to construct tailored resource allocation strategies.

After the 1995 Tokyo subway station sarin gas attacks, 73.9% of the individuals visiting healthcare professionals with complaints on post-exposure symptoms did not require any medical treatment [20]. Our results suggest that, following the initial period of large uncertainty in the disease spread, the magnitude of genuinely affected populations remains significantly larger than the worried-well populations – irrespective of whether the behavior of worried-well individuals is extroverted or introverted. Hence, a conservative policy of preserving at least a cache of critical medical resources in the very early period of the outbreak is advisable. This suggestion is also backed by noting that both asymptomatic testing and vaccines would likely be available as the outbreak progresses.

Finally, our analysis shows that mitigating surges of such populations is important both for reducing spread of the pathogen-based disease as well as reducing the pressure faced by policymakers from such populations. Doing so requires a varied approach from policymakers with effective communication strategies catered for all individuals (rather than ignoring the “small” fraction of worried-well individuals). However, at the most basic level, this requires building trust in institutional services [47]; it is then that public health campaigns are most effective.

## Data Availability

All relevant data are within the manuscript and its Supporting Information files.

## Supporting information

### S1 Appendix

In this appendix we provide additional plots complementing those in the main text: Fig. 4 is analogous to Fig. 2 of the main text while Fig. 5 is analogous to Fig. 3 of the main text. Specifically, we consider two cases, distinguished by extremes of the initial cases, where the initially worried population is ten times less, or more, than the initially infected population.

## Acknowledgments

Bismark Singh is supported by the Deutsche Forschungsgemeinschaft (DFG) - Project number 490948220 titled “Optimal decision making during pandemics”. Both authors thank the COST (European Cooperation in Science and Technology) Action CA22137 Short Term Scientific Mission (STSM) titled “Optimal control strategies in a random two-disease epidemiological model”.

